# Integrating Machine Learning Pipelines for Multimodal Biomarker Prediction in Alzheimer’s and Parkinson’s Disease: A Component of the Neurodiagnoses Framework

**DOI:** 10.1101/2025.08.13.25333561

**Authors:** Nick Osaghae, Manuel Menendez Gonzalez

## Abstract

Alzheimer’s and Parkinson’s diseases are age-related neurodegenerative diseases that often require invasive procedures for diagnosis. Traditional diagnostic methods may fail to capture the interplay between genetic, molecular, and neuroanatomical markers. This manuscript aims to develop interpretable machine learning models that can predict key biomarkers, such as pTau, tTau, Aβ positivity, and motor symptom severity, using non-invasive data. Machine learning models (Random Forest, XGBoost) were trained using ADNI and PPMI baseline data. Using the *APOE4* genotype, MRI volumes, cognitive scores, and demographics as inputs, SHAP was employed to enhance model interpretability. Models achieved AUCs of 0.859 (tTau) and 0.852 (pTau) with recall > 80%. The PD motor severity yielded an MAE of 5.72 and an R^2^ of 0.586. SHAP confirmed the contributions of *APOE4* status, hippocampal atrophy, and dopaminergic asymmetries. The pipelines provide clinically meaningful predictions of biomarker status and motor symptoms, supporting interpretable, multi-axis neurodiagnostic tools within the neurodiagnoses framework.

## INTRODUCTION

Neurodegenerative diseases, most especially Alzheimer’s disease (AD) and Parkinson’s disease (PD), are the two most common brain disorders and leading causes of cognitive decline and disability worldwide. They are primarily characterized by the accumulation of misfolded proteins, such as β-amyloid plaques and tau-containing neurofibrillary lesions in AD and α-synuclein-containing Lewy bodies in PD (1). Despite decades of research, early and accurate diagnosis remains a challenge due to the heterogeneity of symptoms, overlapping clinical presentations, and complexity of biomarkers. This is even more challenging during the prodromal stage of the disease, when interventions are most likely to be effective (2). Artificial Intelligence (AI), especially machine learning (ML), can play a pivotal role in overcoming many of these diagnostic challenges by detecting hidden patterns across multimodal datasets. Traditional diagnostic approaches often rely on isolated clinical or imaging features, which frequently lead to late-stage detection, typically after irreversible neuronal damage has already occurred. However, machine learning models can integrate multiple biomarker sources, including neuroimaging, molecular assays, clinical data, and genetic risk factors, to enhance early detection and explore potential curative benefits (3). The multimodal approach is emerging as a powerful strategy for characterizing the neurodegenerative process. Biomarkers such as structural MRI volumes, cerebrospinal fluid (CSF) levels of Aβ and tau, dopamine transporter (DAT) imaging, *APOE* genotype, and clinical scores have shown complementary value in disease characterization and prediction, even before symptoms appear (4). Building on this, Menéndez et al. (2025) proposed a tridimensional diagnostic framework that combines etiology, molecular pathology, and neuroanatomical-clinical correlation to support earlier and more accurate diagnosis of central nervous system (CNS) disorders (5). The neurodiagnoses framework was developed to operate on this vision. It is a modular, AI-powered system designed for probabilistic modeling, multimodal data integration, and disease progression prediction in complex neurological conditions. The framework leverages interpretable ML models and clinical reasoning to close the gap between these complex data and actionable clinical insights. This study is a component of the neurodiagnoses project, in which we developed two interpretable machine learning pipelines. The first aim is to predict CSF biomarker positivity (Aβ, p-Tau, t-Tau) in Alzheimer’s disease using structural MRI, *APOE4* genotype, and clinical variables from the ADNI dataset. The second aim is to predict the severity of motor score (Unified Parkinson’s Disease Rating Scale, UPDRS III) in Parkinson’s disease using DAT-SPECT imaging, structural MRI, and clinical features from the PPMI dataset. These models highlight stratification based on disease severity and risk, aiming to enhance the triage of clinical trial patients using noninvasive biomarker inputs.

## MATERIALS AND METHODS

### Datasets

We used the baseline data from two publicly available longitudinal neuroimaging and biomarker studies: the Alzheimer’s Disease Neuroimaging Initiative (ADNI) and the Parkinson’s Progression Markers Initiative (PPMI).

### ADNI Pipeline (Classification Tasks)

The ADNI cohort collects multimodal data, including MRI, CSF biomarkers, genetics, and cognitive scores, to study progression from normal aging to Alzheimer’s disease. ADNI data are available upon request approval at adni.loni.usc.edu.(6) We used data from *the ADNIMERGE* composite files with 16,421 and retained only the baseline data entries with CSF Aβ, p-Tau, and t-Tau values (n ≈ 1,200). We focused on predicting (CSF) biomarker positivity of three targets: Aβ positivity: Aβ < 880 pg/mL (exploratory), t-tau/Aβ ratio: > 0.27 (clinical trial triage), and p-tau/Aβ ratio: > 0.023 (clinical trial triage). Input variables included demographic information (age, sex, *APOE4* allele count), MRI-derived regional volumes normalized by intracranial volume (ventricles, hippocampus, entorhinal cortex, fusiform, and middle temporal gyrus), and clinical and cognitive scores (MMSE, MoCA, ADAS, FAQ, CDR-SB, LDELTOTAL, TRABSCORE, and RAVLT). Additional feature-engineered interactions were incorporated to capture domain-specific relationships, such as *APOE4* × age and ADAS13 × ventricular volume.

### PPMI Pipeline (Regression Tasks)

The PPMI cohort aims to identify biomarkers of disease progression in Parkinson’s disease. Like ADNI, it is a publicly available dataset that can be accessed upon request for approval at ppmi-info.org. Data from multiple domains, including DAT-SPECT imaging, MRI, demographics, genotypes, and motor scores, were merged across their CSVs using their unique Participant ID (PATNO). The baseline data across domains, after preprocessing, is approximately 1,716. The target variable is the Unified Parkinson’s Disease Rating Scale (UPDRS-III) motor score (NP3TOT), indicating motor symptom severity. Input features include demographic variables (age, sex), genetic markers (*APOE4* allele count), DAT-SPECT binding ratios (caudate, putamen, and striatum), structural MRI volumes (hippocampus, ventricles, and selected cortical metrics), and the Hoehn and Yahr (H&Y) staging score for exploratory analysis.

### Preprocessing

We employed slightly different preprocessing steps for the two pipelines due to the difference in data structures, target variables, and missing patterns. Missing data was handled using different strategies based on the extent of missingness. Features with more than 50% missingness were removed, and variables with low missing rates were imputed with mean or mode imputation in the ADNI pipeline and multivariate imputation by chained equation (MICE) in the PPMI pipeline to capture inter-feature relationships. MRI-derived volumes were normalized by intracranial volume (ICV) to account for individual brain size differences. Categorical variables such as sex and *APOE4* allele status were encoded numerically. We applied feature engineering to both pipelines to enhance model performance with domain knowledge. For ADNI, we created interaction terms such as *APOE4* ^*^ age and ADAS13 ^*^ ventricular volume to capture the combined effects of genetic risks and structural atrophy. In the PPMI pipeline, striatal asymmetry indices were computed from DAT-SPECT binding ratios, and hemispheric averages were derived for structural MRI metrics to reduce redundancy (7).

### Modelling

#### ADNI Pipeline (Classification)

The objective of the ADNI classification pipeline was to classify individuals on CSF biomarker positivity (Aβ, t-tau/Aβ, and p-tau/Aβ). We tested different models to identify the best performing, starting with logistic regression as a baseline and followed by more advanced ensemble models, Random Forest, XGBoost, and LightGBM. We trained the models incrementally, adding feature engineering and hyperparameter tuning to optimize performance. Random Forest tuning was performed using five-fold cross-validation, adjusting parameters such as the number of trees (n_estimators), tree depth (max_depth), minimum samples per split (min_samples_split), maximum features (max_features), and complexity pruning (ccp_alpha). The slight class imbalance observed (positive-to-negative ratio ~1.15:1) was handled using class weight scaling to mitigate bias. We evaluated the models with standard classification evaluation metrics like Receiver Operating Characteristic - Area Under the Curve (ROC-AUC) and Recall, etc. In clinical trial recruitment and early diagnostic triage, the main objective is to minimize as much as possible the missed detections of biomarker-positive individuals, as these patients represent the critical target population for therapeutic intervention and research enrollment. This is why we prioritized recall (sensitivity) and area under the ROC curve (AUC) as primary performance metrics. High recall ensures that most of the true positives are captured, reducing the risk of excluding eligible patients from trials. AUC provides a robust, threshold-independent measure of model discrimination across the varying clinical cutoffs, supporting generalizability. We used Shapley Additive explanation (SHAP) to assess feature contributions and transparency for the models.

#### PPMI Pipeline (Regression)

The PPMI pipeline aims to predict motor severity using the Unified Parkinson’s Disease Rating Scale (UPDRS-III total score) as a continuous target. We adopted a similar strategy to compare and identify promising models, as in the ADNI pipeline, starting with linear regression and extending to Random Forest, XGBoost, LightGBM, and a shallow artificial neural network (ANN). We used Optuna hyperparameter tuning to optimize the ensemble models with five-fold cross-validation. To reduce dimensionality and increase generalizability, we applied Recursive Feature Elimination with Cross-Validation (RFECV), which selected the most informative features that were used in the final model training. We explored the effect of adding the Hoehn and Yahr score to the model. The models in the regression pipeline were evaluated with standard regression evaluation metrics: MAE, RMSE, and R^2^, with SHAP interpretation.

#### Integration into the Tridimensional Annotation Framework

The pipeline was integrated into the neurodiagnoses framework, which conceptualizes neurodegenerative diseases along three axes: [1] Etiology: genetic and demographic risk factors (e.g., *APOE4*, age), [2] Molecular Markers: CSF Aβ, tau, DAT-SPECT, and [3] Clinical-anatomical: MRI-derived structures, cognitive, and motor scores (e.g., hippocampus, ventricles, MoCA, LDETOTAL, TRABSCORE, RAVLT). We conceived that the prediction of CSF biomarker positivity can form part of the molecular marker axis, while motor scores can contribute to diagnosis on the clinical-anatomical side. This system supports a probabilistic and interpretable approach to the diagnosis of neurodegenerative diseases, building a foundation for early diagnosis, patient stratification in clinical trials, and precision medicine.

## RESULTS

We developed supervised machine learning models for two predictive pipelines: (1) ADNI classification tasks for predicting CSF biomarker positivity (Aβ42, Tau, and pTau) using noninvasive features (structural MRI volume, *APOE4* genotype, and cognitive scores) and (2) PPMI regression tasks for predicting motor score severity in Parkinson’s disease measured by the Unified Parkinson’s Disease Rating Scale Part III (UPDRS-III) using again noninvasive imaging and demographic features. Model performance was evaluated using clinically relevant metrics: AUC, recall, etc., for ADNI classification tasks; and MAE, RMSE, and R^2^ for the PPMI regression tasks. We also assessed the feature importance weight and its directional impact in the models.

### AD Biomarker Prediction (ADNI)

#### Aβ42 Positivity

Amyloid pathology represents the earliest detectable stage in Alzheimer’s disease. We conducted an exploratory classification of Aβ42 positivity (Aβ42 < 880 pg/ml) in conjunction with *APOE4* status, MRI volumes, and clinical scores, achieving moderate predictive accuracy with XGBoost, which yielded the best performance (AUC = 0.81, recall = 80.3%, precision = 71.8%).

#### pTau Positivity

The prediction of pTau/Aβ42 positivity (ratio > 0.023) demonstrated high discriminative performance, with Random Forest achieving the best metrics. (AUC = 0.851, recall = 81%, and specificity = 70%);Table 1). This performance aligns with the sensitivity requirements for clinical trial screening where minimizing false negatives is critical (8). While logistic regression showed slightly higher sensitivity (recall = 87%) and AUC (0.859), this came at the cost of reduced precision, potentially increasing false positives in downstream screening. Random Forest provided comparable sensitivity while improving precision and maintaining interpretability and robustness.

**Table 1:** Performance comparison across models for pTau positivity. Random Forest offered the best balance between recall and precision

| Model | AUC-ROC | Recall (1) | Precision (1) | F1-Score (1) |
| --- | --- | --- | --- | --- |
| Logistic Regression | 0.859 | 0.87 | 0.77 | 0.82 |
| Optimized Logistic Regression | 0.855 | 0.73 | 0.86 | 0.79 |
| Random Forest (Best) | 0.851 | 0.81 | 0.78 | 0.79 |
| XGboost | 0.838 | 0.80 | 0.75 | 0.77 |

#### tTau Positivity

For the prediction of tTau/Aβ42 positivity (ratio > 0.27), XGBoost demonstrated superior performance (AUC = 0.859, recall = 87%, specificity = 70%) compared to other models (Table 2). This indicates a reliable identification of tau-positive individuals, making it suitable for clinical triage applications.

**Table 2:** Model comparison in tTau positivity classification; XGBoost showed the best performance.

| Model | AUC-ROC | Recall (1) | Precision (1) | F1-Score (1) |
| --- | --- | --- | --- | --- |
| Logistic Regression | 0.846 | 0.74 | 0.83 | 0.78 |
| Random Forest | 0.849 | 0.72 | 0.80 | 0.76 |
| XGBoost (Best) | 0.859 | 0.87 | 0.77 | 0.82 |

#### PD Motor Progression Prediction

The regression model’s goal is to predict UPDRS-III motor scores using multimodal baseline features of DAT-SPECT striatal binding ratios, structural MRI volumes, and demographic features like age and *APOE4* status. Among the models evaluated, Random Forest achieved superior performance without Hoehn & Yahr (H&Y) staging (R^2^ = 0.586, MAE = 5.72, RMSE = 7.48), outperforming linear regression and XGBoost (Table 3.3). Including the H&Y feature improved performance significantly (R^2^ = 0.680, MAE = 4.63, RMSE = 6.57), but SHAP revealed that H&Y became the dominant driver, possibly overshadowing the contributions of other biological signals. This is a known phenomenon in multimodal PD studies (9).

**Table 3:** Performance of UPDRS-III prediction models on PPMI data. Random Forest showed the best performance without H&Y

| Model | MAE | MSE | RMSE | R2 |
| --- | --- | --- | --- | --- |
| Linear Regression | 9.75 | 142.86 | 11.95 | 0.035 |
| Random Forest (Best) | 5.72 | 55.96 | 7.48 | 0.59 |
| XGBoost | 6.53 | 76.95 | 8.77 | 0.48 |
| LightGBM | 6.54 | 74.711 | 8.64 | 0.49 |
| Shallow ANN | 6.98 | - | - | 0.41 |
| Random Forest | 4.63 | 43.21 | 6.57 | 0.68 |

#### Model Interpretability and Key Biomarkers with SHAP

To ensure transparency and clinical relevance, we applied SHAP analysis to interpret the final machine learning models and present the contributions of each variable to the prediction. This approach provides both a global explanation through an aggregated feature importance ranking and a local explanation through SHAP beeswarm plots showing how individual features influence prediction. Figure 2 illustrates the SHAP beeswarm plot and feature importance summary for the best-performing model: classification of tTau positivity. This model highlighted the interactive effect of *APOE4* status and age, particularly among individuals over 65, as the most important predictor of tau positivity. Other top predictors included hippocampal volume, ADAS13, MoCA, and FAQ, reflecting genetic, structural, and cognitive decline associated with tau pathology. These findings align with previous literature showing that *APOE4*, neurodegeneration in medial temporal structures, and cognitive dysfunction are predictive of CSF tau elevation (10). We conducted similar SHAP analyses across pTau positivity (*Supplementary Figure SI*); the *APOE4* ^*^ age interaction again emerged as a primary contributor, along with LDELTOTAL and Trail Making Test Part B (TRABSCORE), indicating the role of memory and executive dysfunction in pTau accumulation. Hippocampal atrophy and functional decline (FAQ) also ranked highly (11). For amyloid (Aβ) positivity (*Supplementary Figure S2*), we noticed a similar trend of *APOE4* and age driving prediction, though ventricular volume, mid_temp, hippocampal volume, and ADAS13 scores also contributed; this emphasizes the model’s sensitivity to early-stage neurodegenerative biomarkers (12). In the PD motor progression regression task (*Supplementary Figure S3*), SHAP analysis for Random Forest trained without H&Y identified DAT-SPECT imaging features, especially putamen-total binding ratios and striatal asymmetry, as the most influential feature, followed by age. Noticeably lower DAT-SPECT features like putamen total and caudate drive SHAP values, indicating that lower DAT density is linked with cognitive impairment. This is consistent with well-known patterns of dopaminergic loss and age-related motor decline in Parkinson’s disease. SHAP analysis revealed that H&Y became the dominant contributor to the models when added, masking underlying biological signals; this is a limitation that has been previously observed in PD studies (13). SHAP results indicated that the models are indeed biologically plausible and that they have some potential for clinical utility. While clinical scores carry strong predictive power, their interactions with genetic and neuroimaging markers, especially *APOE4* ^*^ age, provide interpretable and disease-related insights for trial stratification and early diagnosis.

**Figure 1:**
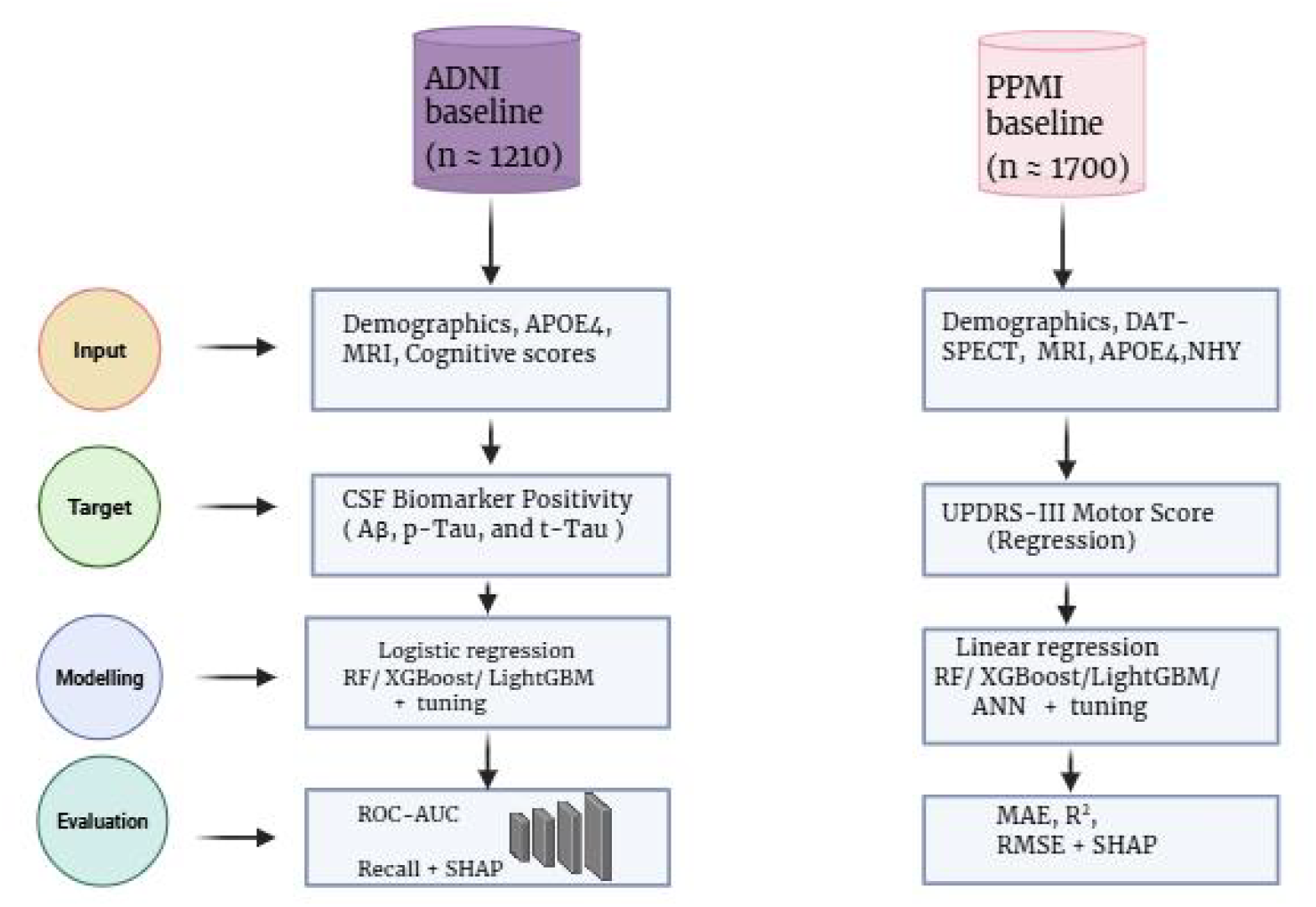
Overview of the data processing and modeling pipeline for the ADNI and PPMI cohort

**Figure 2:**
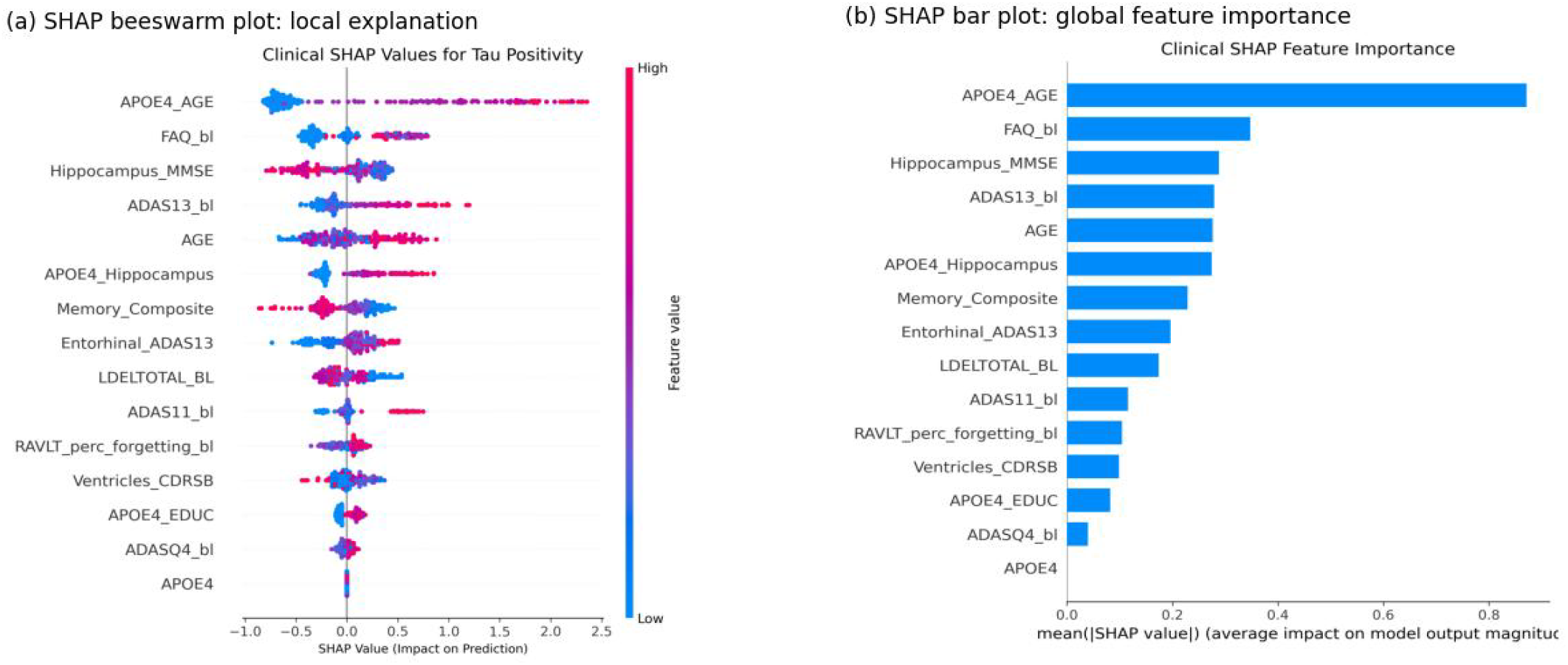
SHAP interpretability of the tTau positivity classification model. (a) Shows the SHAP beeswarm plot with the local explanation of individual predictions (b) SHAP bar plot of the mean absolute values of the global feature importance

## DISCUSSION

This study aimed to present interpretable machine learning pipelines for predicting cerebrospinal fluid biomarker positivity (Aβ, tTau, and pTau) in Alzheimer’s disease using baseline non-invasive data from ADNI and motor symptom severity in Parkinson’s disease (PD) using baseline imaging and clinical features from the PPMI cohort. The main goal was to assess whether accessible, multimodal biomarkers (e.g., MRI-derived volumes, DAT-SPECT, *APOE4* genotype, demographics, cognitive scores) can serve as viable surrogates for more invasive or costly diagnostic methods such as lumbar punctures or PET imaging, with the potential to enhance early detection, risk stratification, and clinical trial enrollment as part of the objectives of the neurodiagnoses framework. In terms of clinical relevance, our results show that the use of noninvasive biomarkers in approximating CSF status is feasible. In the Alzheimer’s disease models, XGBoost and Random Forest yielded AUC values of 0.859 for tTau positivity and 0.852 for pTau, respectively, with both having recall values exceeding 80%. These models not only surpassed suggested minimal thresholds for clinical utility but also maintained high sensitivity in identifying biomarker-positive individuals. The *APOE4* genotype and its interaction with age were important predictors that consistently influenced output across all CSF biomarker models, especially in individuals above 65; this confirms the established association of the *APOE4* genotype and age-related neurodegeneration with amyloid and tau pathology (14). Hippocampal volume and entorhinal atrophy are known early neurodegenerative biomarkers that also ranked highly in SHAP analysis, reinforcing their diagnostic relevance and the contribution of multiple biomarkers (15). These findings suggest that noninvasive features can robustly approximate the presence of CSF biomarkers, reducing reliance on invasive methods such as lumbar puncture and enhancing accessibility in low-resource environments or supporting potential utility in early screening in clinical trials. In the Parkinson’s disease motor severity (UPDRS-III) regression model, the Random Forest model achieved promising performance with baseline data, reaching an R^2^ of 0.586 and a mean absolute error (MAE) of 5.72. Surpassing MRI-only models and approaching a clinically relevant error margin (3.25-5.55) (16) (17). DAT-SPECT features, particularly those of putamen and caudate asymmetry, were the most influential predictors. These findings align very well with established patterns of dopaminergic degeneration in PD and support the use of neuroimaging biomarkers in predicting motor progression (18)(13). Performance improved sharply with Hoehn and Yahn (H&Y) scores (R^2^ = 0.68), but SHAP analysis revealed that this improvement is mainly contributed by H&Y; this highlights the risk of label leakage, as clinical scores that strongly correlate with the outcome can artificially inflate performance while reducing the effect of biological signals. It is important to avoid proximal clinical features to ensure unbiased predictions. Beyond model evaluation and performance, this work contributes to the neurodiagnoses framework that aims to create a robust diagnostic system that conceptualizes neurodegenerative disease across multiple diagnostic axes (etiology, molecular markers, and clinical-anatomical). The best-performing pipeline for tTau positivity has been successfully integrated into Axis 2 (Molecular markers) of the framework as its primary decision engine. The model now supports real-time annotation of molecular pathology. Some limitations must be acknowledged. Firstly, this study relied on baseline data only, therefore losing its ability to model longitudinal disease progression across timelines. Secondly, the study used ADNI and PPMI datasets; these are standardized datasets that may not reflect the variability of data available in real-world clinical practice. Also, thresholds for CSF biomarker positivity were cohort-specific for the ADNI cohort, and they may not generalize across sites or populations. Future work should incorporate longitudinal features and time series modelling. Time-dependent features may improve early detection and support prognostic applications. Future work should also incorporate federated learning to ensure generalizability across diverse populations and institutional settings.

## CONCLUSION

This study demonstrates that machine learning models trained on multimodal, noninvasive baseline data can predict CSF biomarker positivity in Alzheimer’s disease and motor symptom severity in Parkinson’s disease with clinically meaningful accuracy. The tTau and pTau models exceeded minimal thresholds for clinical utility. The models offered interpretable outputs and showed important signals, with *APOE4*-age interactions and MRI-derived volumes emerging as biologically grounded predictors.

The study supported a successful integration of the tTau model into the Neurodiagnoses framework, powering molecular-level inference (Axis 2) as part of a 3-axis diagnostic map. This integration supports the framework’s vision to provide clinicians with transparent modular AI tools that complement expert decision-making and reduce diagnostic burden.

## Data Availability

All data produced in the present work are contained in the manuscript

https://www.ppmi-info.org/access-data-specimens/download-data

https://adni.loni.usc.edu/data-samples/adni-data/

## SUPPLEMENTARY INFORMATION

**Supplementary figure S1:**
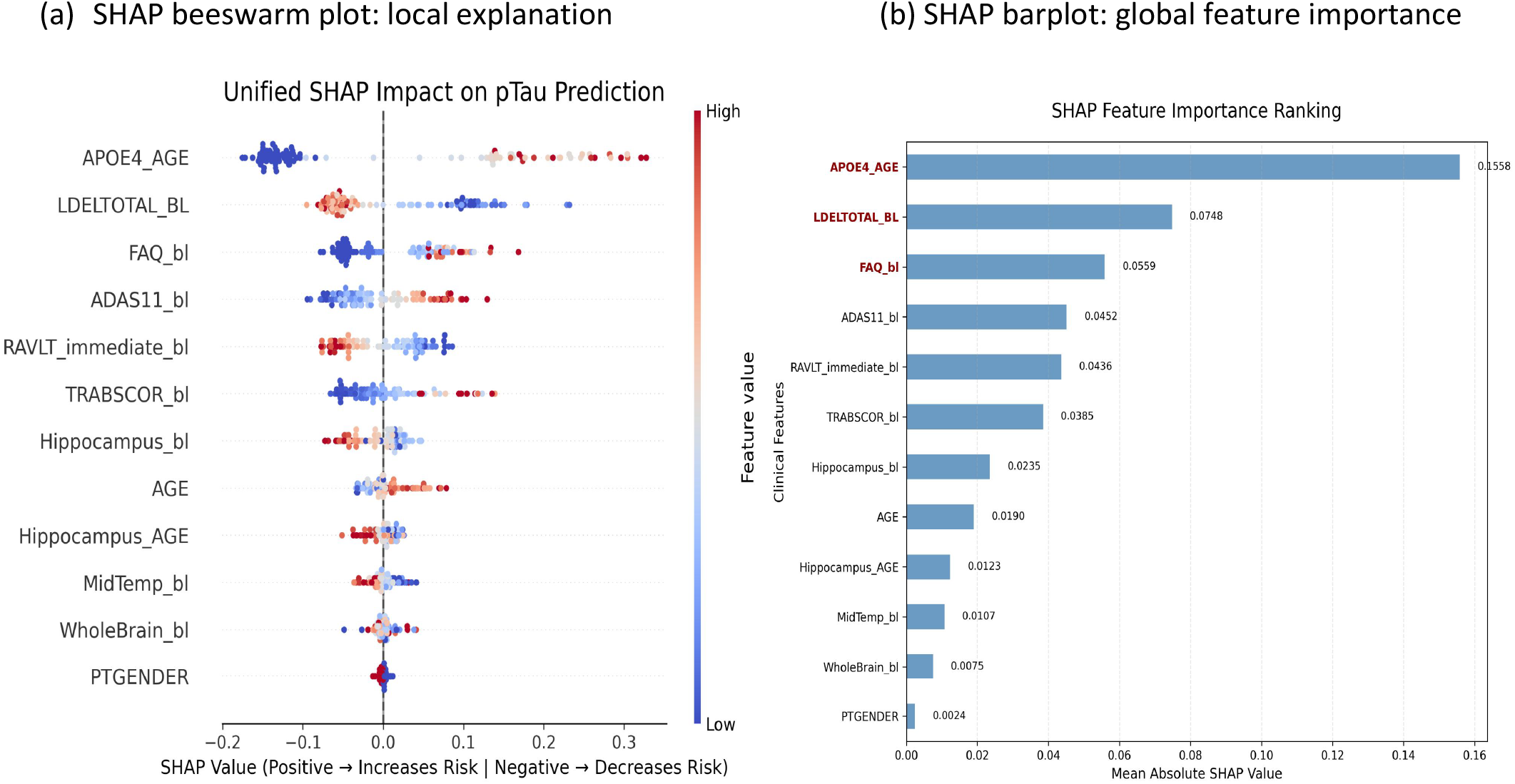
SHAP interpretability of the pTau positivity classification model. (a) Shows the SHAP beeswarm plot with the local explanation of individual predictions. (b) SHAP barplot of the mean absolute values of the global feature importance

**Supplementary figure S2:**
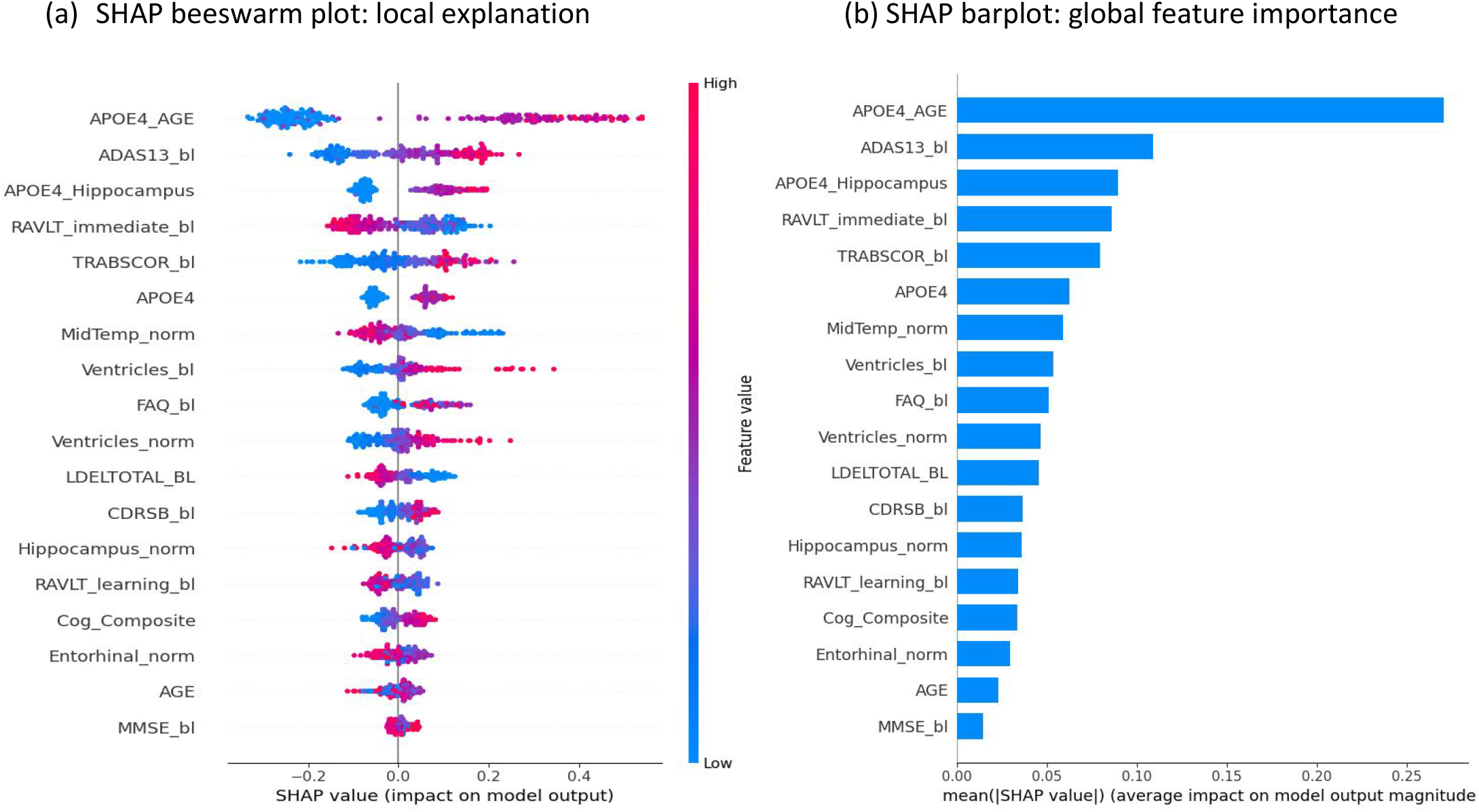
SHAP interpretability of the amyloid (Aβ) positivity classification model. (a) HAP beeswarm plot with the local explanation of individual predictions. (b) SHAP barplot of the mean absolute values of the global feature importance

**Supplementary figure S3:**
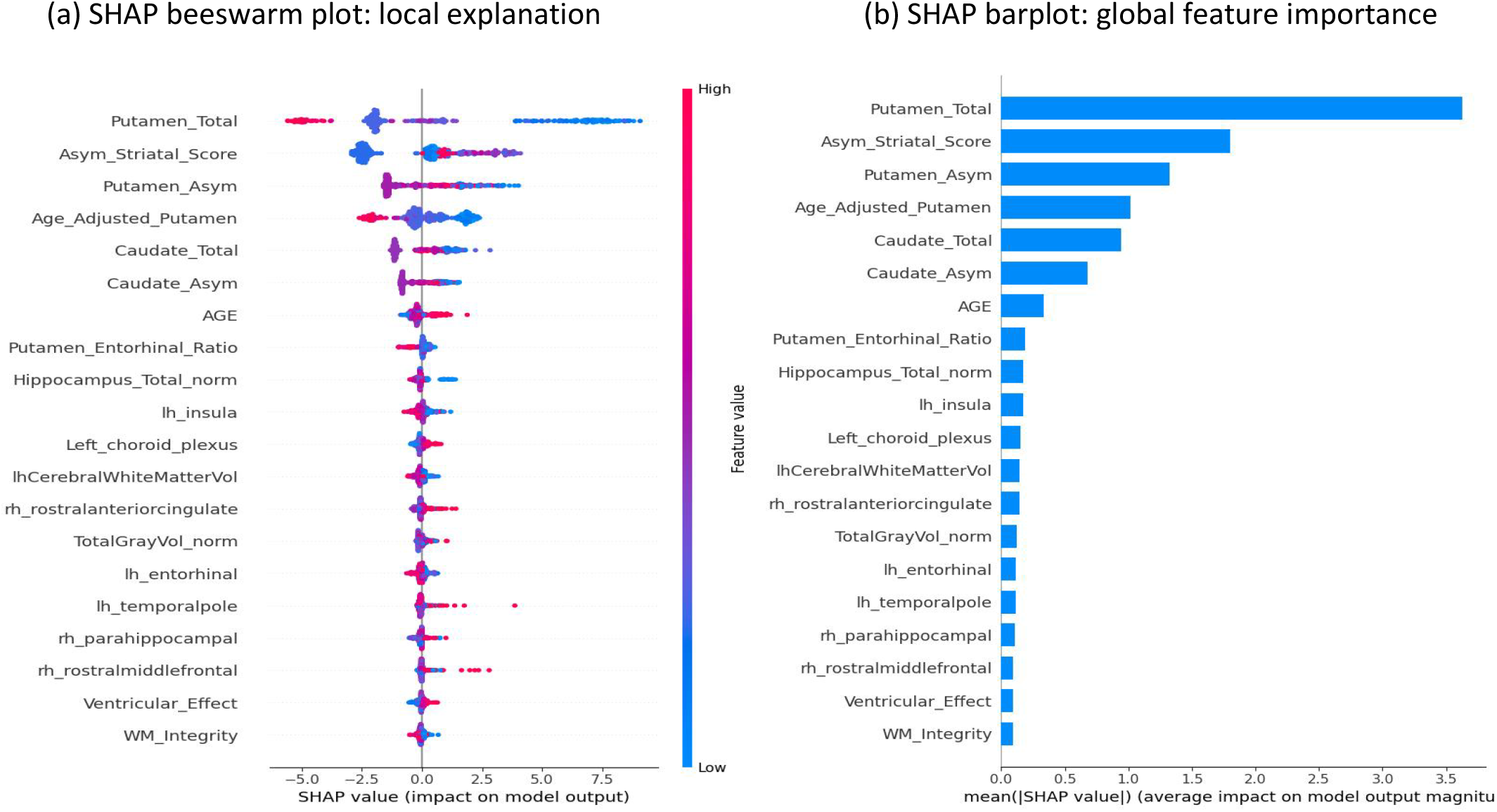
SHAP interpretability of the PD motor regression model. (a) Shows the SHAP beeswarm plot with the local explanation of individual predictions. (b) SHAP barplot of the mean absolute values of the global feature importance

